# Immunogenicity and durability against Omicron BA.1, BA.2 and BA.4/5 variants at 3 to 4 months after a heterologous COVID-19 booster vaccine in healthy adults with a two-doses CoronaVac vaccination

**DOI:** 10.1101/2022.11.24.22282735

**Authors:** Suvichada Assawakosri, Sitthichai Kanokudom, Nungruthai Suntronwong, Jira Chansaenroj, Chompoonut Auphimai, Pornjarim Nilyanimit, Preeyaporn Vichaiwattana, Thanunrat Thongmee, Thaneeya Duangchinda, Warangkana Chantima, Pattarakul Pakchotanon, Donchida Srimuan, Thaksaporn Thatsanatorn, Sirapa Klinfueng, Natthinee Sudhinaraset, Nasamon Wanlapakorn, Juthathip Mongkolsapaya, Sittisak Honsawek, Yong Poovorawan

**Affiliations:** Center of Excellence in Clinical Virology, Faculty of Medicine, Chulalongkorn University, Bangkok, 10330, Thailand; Center of Excellence in Osteoarthritis and Musculoskeleton, Faculty of Medicine, Chulalongkorn University, King Chulalongkorn Memorial Hospital, Thai Red Cross Society, Bangkok, 10330, Thailand; Molecular Biology of Dengue and Flaviviruses Research Team, National Center for Genetic Engineering and Biotechnology, National Science and Development Agency, NSTDA, Pathum Thani 12120, Thailand; Division of Dengue Hemorrhagic Fever Research, Faculty of Medicine, Siriraj Hospital, Mahidol University, Bangkok 10700, Thailand; Siriraj Center of Research Excellence in Dengue and Emerging Pathogens, Faculty of Medicine, Siriraj Hospital, Mahidol University, Bangkok 10700, Thailand; Wellcome Centre for Human Genetics, Nuffield Department of Medicine, University of Oxford, Oxford, OX3 7BN, UK; Chinese Academy of Medical Science (CAMS) Oxford Institute (COI), University of Oxford, Oxford, UK; FRS(T), the Royal Society of Thailand, Sanam Sueapa, Dusit, Bangkok 10330, Thailand

**Keywords:** COVID-19 vaccine, durability, CoronaVac, heterologous booster, neutralizing antibody, Omicron

## Abstract

**Objectives:** Several countries have authorized a booster vaccine campaign to combat the spread of COVID-19. Data on persistence of booster vaccine-induced immunity against new Omicron subvariants are still limited. Therefore, our study aimed to determine the serological immune response of COVID-19 booster after CoronaVac-priming.

**Methods:** A total of 187 CoronaVac-primed participants were enrolled and received an inactivated (BBIBP), viral vector (AZD1222) or mRNA vaccine (full-/half-dose BNT162B2, full-/half-dose mRNA-1273) as a booster dose. The persistence of humoral immunity both binding and neutralizing antibodies against wild-type and Omicron was determined on day 90– 120 after booster.

**Results:** A waning of total RBD immunoglobulin (Ig) levels, anti-RBD IgG, and neutralizing antibodies against Omicron BA.1, BA.2, and BA.4/5 variants was observed 90–120 days after booster vaccination. Participants who received mRNA-1273 had the highest persistence of the immunogenicity response, followed by BNT162b2, AZD1222, and BBIBP-CorV. The responses between full and half doses of mRNA-1273 were comparable. The percentage reduction of binding antibody ranged from 50% to 75% among all booster vaccine.

**Conclusions:** The antibody response substantially waned after 90–120 days post-booster dose. The heterologous mRNA and the viral vector booster demonstrated higher detectable rate of humoral immune responses against the Omicron variant compared to the inactivated BBIBP booster. Nevertheless, an additional fourth dose is recommended to maintain immune response against infection.

**Highlights:**

- The persistence of antibody responses is different among three vaccine platforms.
- Highly remained antibody levels were observed with the mRNA and viral vector booster.
- The half-dose mRNA-1273 can be used interchangeably with the full-dose mRNA-1273.
- The neutralizing activity against BA.5 was lower than wild type and BA.2 subvariant.
- A fourth dose is recommended for individuals who received an inactivated booster.

## 1. Introduction

With the ongoing coronavirus disease 2019 (COVID-19) pandemic, the emerging Omicron variant of severe acute respiratory syndrome coronavirus 2 (SARS-CoV-2) is considered more transmissible than other previous variants of concern, including Alpha, Beta, and Delta variants (Kumar et al., 2022). Therefore, vaccination is an essential tool to alleviate the spread of the COVID-19 pandemic. The inactivated virus vaccine platform is one of the most widely used as primary regimen among all COVID-19 vaccines. CoronaVac (Sinovac Biotech Co., Ltd., Beijing, China) is one of a whole inactivated virus COVID-19 vaccines that have been approved for use in more than 56 low- and middle-income countries around the world, including Thailand (COVID-19 tracker, 2022). Nevertheless, several studies have demonstrated that the neutralization of the Omicron variant was highly limited in sera from those who received the primary series of COVID-19 vaccines, for either the mRNA or viral vector or inactivated platforms (Edara et al., 2022, Lu et al., 2022, Muik et al., 2022, Planas et al., 2022). Therefore, the use of third dose COVID-19 vaccines has been recommended by the US Food and Drug Administration to combat the new Omicron variant (Centers for Disease Control and Prevention, 2022).

In late December 2021, a mass booster dose vaccination campaign was first recommended in Thailand by the Ministry of Public Health, especially for adults previously vaccinated with two doses of CoronaVac. Recently, we reported the results of a third dose booster study in healthy adults previously primed with two doses of CoronaVac. Our finding showed that a third heterologous dose elicited a robust immune response against the Omicron variant (Assawakosri et al., 2022b, Kanokudom et al., 2022b). Moreover, a booster with BNT162b2 in CoronaVac-primed individuals improved vaccine effectiveness (VE) to 92.7% for protection against COVID-19 infection in Brazil (Cerqueira-Silva et al., 2022b). A significant challenge in controlling the COVID-19 pandemic is the waning of vaccine-induced immunity. A previous COV-boost study reported the decline in immunogenicity three months after the third dose using viral vector platforms, AZD1222 (AstraZeneca, Oxford, UK) (Liu et al., 2022). However, it is currently unclear on immunogenicity beyond one month following the third heterologous dose in CoronaVac-primed individuals as is how rapidly protection from booster dose wanes over time.

Subvariants of the Omicron are continuously emerging during the COVID-19 pandemic. Initially, Omicron variants were categorized into several descendent subvariants, including BA.1, BA.1.1, BA.2, BA.2.2, and BA.3 (Viana et al., 2022; Yamasoba et al., 2022). Subsequently, two new Omicron subvariants that harbor the L452R spike protein mutation have been designated as BA.4 and BA.5 (Tegally et al., 2022). Both BA.4 and BA.5 have identical spike protein sequences (hereafter BA.4/5) (Zhou et al., 2022). A previous study showed that the BA.4/5 subvariants are more pathogenic than BA.1 and are highly resistant to BA.1 and BA.2-infected sera (Kimura et al., 2022). Furthermore, BA.4/5 exhibited an 18.3-fold higher infectivity rate than that observed in BA.2 (Zhou et al., 2022). However, there are limited data on BA.4/5 immune evasion in third-dose heterologous vaccinated individuals. Therefore, the present study aimed to evaluate the durability of immune protection at three to four months following four different heterologous booster vaccines in adults who completed a two-dose CoronaVac. In addition, we also investigated immunogenicity against the newly emerging Omicron variants BA.2 and BA.4/5 subvariants.

## 2. MATERIALS AND METHODS

### 2.1 Study design and participants

The study protocol was described in our previous studies (Assawakosri et al., 2022b, Kanokudom et al., 2022b). Briefly, this study was a prospective cohort study of heterologous third-dose vaccination. All participants were Thai adults over 18 years of age with no previous or current diagnosis of COVID-19 infection and who had received the CoronaVac 2-dose vaccine (Sinovac Biotech Co., Ltd., Beijing, China) with the 6±1 months interval period. All participants were allocated to receive a booster dose of the COVID-19 vaccine, including BBIBP-CorV, AZD1222, BNT161b2 (full- or half-dose), and mRNA-1273 (full- or half-dose). The study flow is illustrated in Figure 1. This study was conducted in the Clinical Trial Unit, the Center of Excellence in Clinical Virology of Chulalongkorn University, Bangkok, Thailand. The study was approved by the Institutional Review Board of the Faculty of Medicine, Chulalongkorn University (IRB numbers 546/64 and 498/65). The study was registered with the Thai Registry of Clinical Trials (TCTR 20210910002). Written informed consent was obtained from each participant before the enrollment and this cohort study was conducted in accordance with the Declaration of Helsinki.

**Figure 1.**
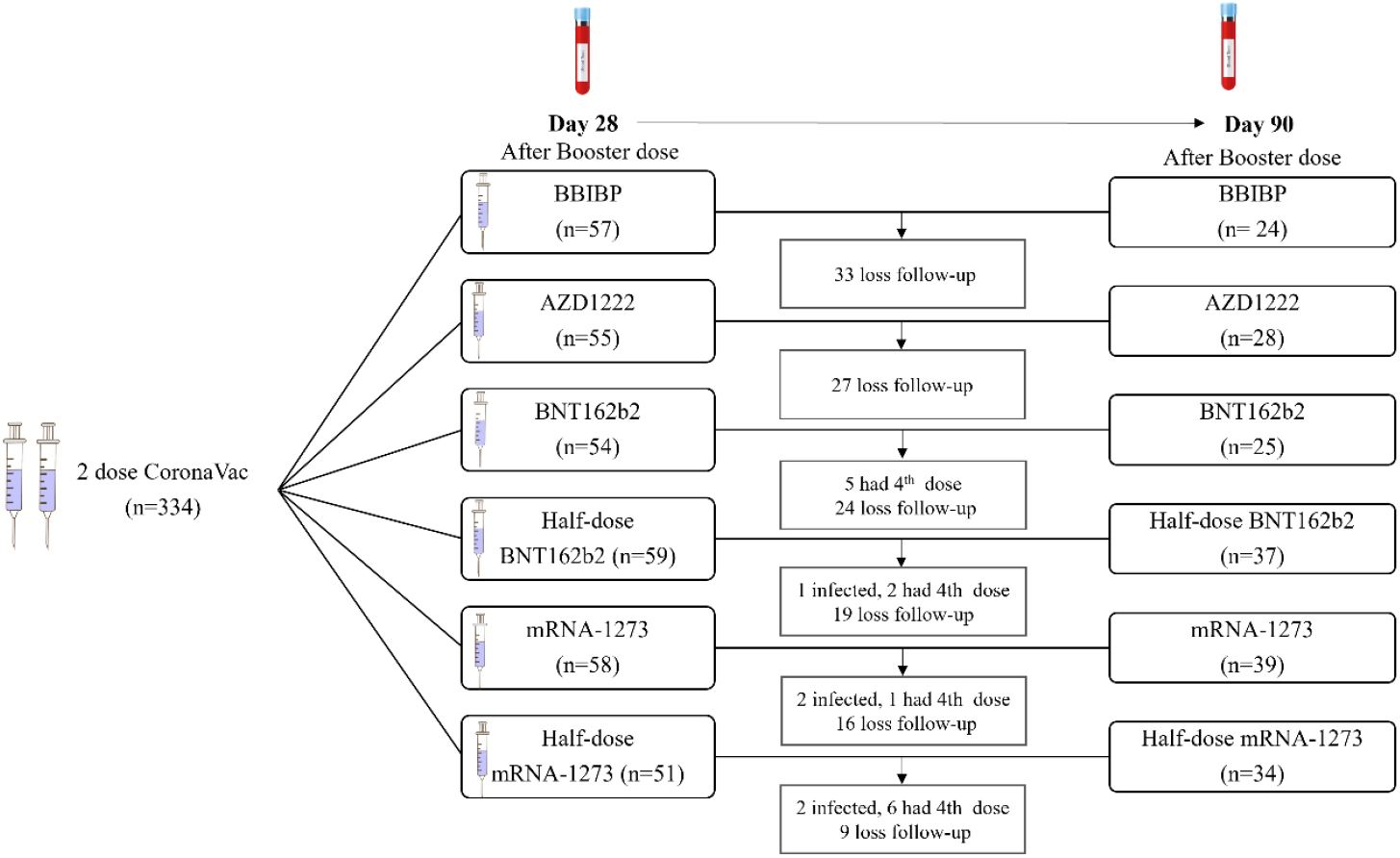
Diagram of recruited participants and study groups. In our previous study, 334 participants who had completed two injections of CoronaVac were enrolled and received a single booster dose of including BBIBP (n=57), AZD1222 (n=55), BNT162b2 (n=54), half-dose BNT162b2 (n=59), mRNA-1273 (n=58), or half-dose mRNA-1273 (n=51) at Day 28 after booster timepoint. This extended study, a total of 187 matched individuals including those receiving BBIBP (n=24), AZD1222 (n=28), BNT162b2 (n=25), half-dose BNT162b2 (n=37), mRNA-1273 (n=39), or half-dose mRNA-1273 (n=34) who completed day 90 to 120 follow up timepoints were eligible for final analysis of binding and neutralizing antibodies.

### 2.2 Study vaccine

The study vaccines were as follows: inactivated: BBIBP-CorV (Sinopharm, Beijing, China), viral vector: AZD1222 (AstraZeneca, Oxford, UK), mRNA: 30 μg BNT162b2 (full-dose group) and 15 μg (half-dose group) (Pfizer-BioNTech Inc., New York City, NY, USA), mRNA: 100 μg mRNA-1273 (full-dose group) (Moderna Inc., Cambridge, MA, USA), and 50 μg mRNA-1273 (half-dose group).

### 2.3 Sample collection

Peripheral venous blood samples (10–15 mL) were collected on day 28, and 90 to 120 after vaccination. The clot blood tubes were then centrifuged at 2000 rpm for 10 minutes at room temperature to collect serum samples. All specimens were stored at -20°C until further analysis.

### 2.4 Total RBD Ig, anti-RBD IgG, and anti-nucleocapsid assay

Serum samples were used to measure total immunoglobulin (Ig) specific for the RBD of SARS-CoV-2 using an electrochemiluminescence immunoassay—Elecsys SARS-CoV-2 S (Roche Diagnostics, Basel, Switzerland)—according to the manufacturer’s instructions and as previously described (Kanokudom et al., 2022). The Ig titer was determined as unit per milliliter (U/mL), and the level ≥ 0.8 U/mL was considered a positive detection.

The levels of IgG specific to an RBD and nucleocapsid protein of the SARS-CoV-2 virus in serum samples were detected by the chemiluminescence microparticle immunoassay (Abbott, Sligo, Ireland). Anti-RBD IgG is a quantitative assay with a limit of detection level at ≥ 21 AU/mL, a lower level of less than 50 AU/mL is considered a negative result. While the Anti-nucleocapsid IgG is a semi-quantitative technique, the level ≥ 1.4 sample/cutoff (S/C) was defined as positive detection.

### 2.5 Surrogate virus neutralization assay for Wuhan and Omicron variants

Neutralizing activities against wild type (Euroimmun, Lubeck, Germany) and a cPassTM SARS-CoV-2 surrogate virus neutralization test (sVNT) (GenScript Biotech, Piscataway, NJ, USA) were used to measure NAb titers against SARS-CoV-2 variants. Recombinant RBD of Omicron; BA.2 strains (G339D, S371F, S373P, S375F, T376A, D405N, R408S, K417N, N440K, S477N, T478K, E484A, Q493R, Q498R, N501Y, Y505H) and BA.4/5 strains (G339D, S371F, S373P, S375F, T376A, D405N, R408S, K417N, N440K, L452R, S477N, T478K, E484A, F486V, Q498R, N501Y, Y505H) were incubated in 96-well plates coated with recombinant human ACE2 and the sera sample of vaccinees. Samples with a percentage inhibition (% inhibition) greater than or equal to 35% threshold for wild type and greater than or equal to 30% for the Omicron BA.2 and BA.4/5 subvariants were considered ‘seropositive’ for SARS-CoV-2 neutralizing antibodies (Kanokudom et al., 2022a).

### 2.6 Focus reduction neutralization test (FRNT50) against the BA.1 and BA.2 variants

Neutralization antibody titers were measured against BA.1 variants (accession number: EPI_ISL_8547017) and BA.2 (EPI_ISL_11698090) of the SARS-CoV-2 Omicron. The Focus reduction neutralization assay was performed as previously described (Assawakosri et al., 2022a, 2022b). Additionally, the 50%focus reduction was calculated, and the half maximum inhibitory concentration (IC50) was determined using PROBIT regression analysis (SPSS Inc., Chicago, IL, USA). The detection limit of the assay is 1:20 and NAb values below the detection limit were substituted with a titer of 10.

### 2.7 Statistical analysis

All statistical analyses were performed using the Statistical Package for the Social Sciences (SPSS) v.22.0 (SPSS Inc., Chicago, IL, USA). Figures were generated using GraphPad Prism v9.0 (GraphPad Software, San Diego, CA, USA). The categorical data comparison, including sex and comorbidities, was performed using Pearson’s chi-square test. Total RBD Ig, anti-RBD IgG, and NAb levels were reported as geometric mean titers (GMT) with a 95% confidence interval (CI). The geometric mean ratio of immunogenicity between day 90–120 and day 28 timepoints (GMR D90–120/28) was calculated using the GMT logarithmically transformed and was performed using a paired *t* test. Analysis of variance (ANOVA) with Bonferroni adjustment or Kruskal–Wallis H test (for nonparametric data) was used to compare independent groups. The percentage inhibition of the surrogate neutralization assay and anti-nucleocapsid (N) IgG was calculated as the median with the interquartile range (IQR). A *P*-value of <0.05 (*), <0.01 (**), <0.001 (***) was considered statistically significant.

## 3. RESULTS

### 3.1 Demographic data

Of the 334 participants vaccinated with a different COVID-19 booster doses from two previous studies between September and November 2021, 187 participants were included in this long-term follow-up study with assessments on day 90 to 120. The demographic characteristics of this study are presented in Table 1. All participants were healthy Thai adults (well-control comorbidities were acceptable) between 20 and 69 years of age. The mean age (range) of participants was 40.8 (20–69), 43.7 (31–64), 45.0 (20–62), 40.8 (22–55), 40.6 (20–63), 36.7 (21– 57), and 40.5 (20–69) for BBIBP, AZD1222, BNT161b2 (full- or half-dose), and mRNA-1273 (full- or half-dose), respectively. The average interval between the second and booster doses was 160.7 (115–237) days. The baseline sex and comorbidities of the participants were comparable among all groups. Whereas the mean age of the mRNA-1273 group was significantly lower than other groups. Age was used for statistical adjustment. During the cohort period, two participants were excluded from the study due to anti-N IgG seroconversion on day 90 to 120 were suspected of COVID-19 infection.

**Table 1.** Demographics and baseline characteristics of the enrolled participants.

|  | **Total** | **BBIBP** | **AZD1222** | **BNT162b2** | **Half**<br>**BNT162b2** | **mRNA-1273** | **Half**<br>**mRNA-1273** |
| --- | --- | --- | --- | --- | --- | --- | --- |
| **Total number (n)** | 187 | 24 | 28 | 25 | 37 | 39 | 34 |
| **Mean age (range) year** | 40.9 (20-69) | 43.7 (31-64) | 45.0 (20-62) | 40.8 (22-55) | 40.6 (20-63) | 37.0 (21-57) | 40.6 (20-69) |
| **Sex** |  |  |  |  |  |  |  |
| **Male (%)** | 88/187<br>(47.1%) | 12/24<br>(50.0%) | 10/28<br>(35.7%) | 17/25<br>(68.0%) | 16/37<br>(43.2%) | 18/39<br>(46.2%) | 15/34<br>(44.1%) |
| **Female (%)** | 99/187<br>(52.9%) | 12/24<br>(50.0%) | 18/28<br>(64.3%) | 8/25<br>(32.0%) | 21/37<br>(56.8%) | 21/39<br>(53.8%) | 19/34<br>(55.9%) |
| **Underlying disease (%)** |  |  |  |  |  |  |  |
| **Allergy** | 12/187 (6.4%) | 1/24 (4.2%) | 2/28 (7.1%) | 1/25 (4.0%) | 2/37 (5.4%) | 3/39 (7.7%) | 3/34 (8.8%) |
| **Cardiovascular** | 3/187 (1.6%) | 1/24 (4.2%) | 2/28 (7.1%) | - | - | - | - |
| **diseases** | 10/187 (5.3%) | 1/24 (4.2%) | 4/28 (14.3%) | 2/25 (8.0%) | 2/37 (5.4%) | 1/39 (2.6%) | - |
| **Diabetes Mellitus** | 3/187 (1.6%) | 1/24 (4.2%) | 1/28 (3.6%) | 1/25 (4.0%) | - | - | - |
| **Dyslipidemia** | 15/187 (8.0%) | 1/24 (4.2%) | 3/28 (10.7%) | 2/25 (8.0%) | 4/37 (10.8%) | 3/39 (7.7%) | 2/34 (5.9%) |
| **Hypertension** | 3/187 (1.6%) | 1/24 (4.2%) | - | - | - | 1/39 (2.6%) | 1/34 (2.9%) |
| **Thyroid** | 10/187 (5.3%) | 2/24 (8.3%) | 1/28 (3.6%) | - | 2/37 (5.4%) | 2/39(5.1%) | 3/34 (8.8%) |
| **Other (Gout, Asthma,**<br>**COPD, etc.)** |  |  |  |  |  |  |  |
| **Interval between the 2<sup>nd</sup> dose**<br>**and the booster dose** | 160.6<br>(115-237) | 166.6<br>(115-197) | 153.6<br>(130-191) | 144.2<br>(120-166) | 162.3<br>(148-213) | 168.5<br>(148-237) | 163.5<br>(150-210) |
| **Follow-up** |  |  |  |  |  |  |  |
| **Third visit** |  |  |  |  |  |  |  |
| **Mean (range) day** | 28.2<br>(22-36) | 29.0<br>(25-35) | 27.9<br>(22-29) | 28.1<br>(28-30) | 28.3<br>(27-30) | 27.7<br>(27-32) | 28.6<br>(28-36) |
| **Fourth visit** |  |  |  |  |  |  |  |
| **Mean (range) day** | 100.1<br>(86-127) | 93.3<br>(89-113) | 120<br>(117-121) | 120.6<br>(119-127) | 92.8<br>(90-106) | 91.5<br>(89-102) | 91.3<br>(86-95) |

### 3.2 Measurement of Total RBD Ig, Anti-RBD IgG, and Anti-N IgG

Overall, after booster dose vaccination, there was a significant reduction in antibody levels, both total RBD Ig and anti-RBD IgG, on day 90 to 120. At 28 days after booster vaccination, the GMT of the total RBD Ig were 1231, 10,766, 21,240, 22,345, 36,845, and 28,087 U/mL for BBIBP, AZD1222, BNT161b2 (full- or half-dose), and mRNA-1273 (full- or half-dose), respectively. Subsequently, the GMT of the total RBD Ig of six groups were decreased to 620, 3984, 5514, 9722, 10,853, and 10,587 U/mL, respectively, at 90–120 days after booster dose. Among the six groups, the decay rate of total RBD Ig levels in the BBIBP group was lower than that in other groups, with a GMR D90–120/28 ratio of 0.50 (95%CI: 0.41–0.61). While the GMR D90–120/28 ratios were comparable in the mRNA booster groups with 0.26 (95%CI: 0.22–0.30) for BNT162b2 and 0.30 (95%CI: 0.27–0.32) for the mRNA-1273 group. Interestingly, a comparable decay rate was evident between half-dose and full-dose mRNA-1273. Similar trends were observed in anti-RBD IgG levels. The percentage reduction of each booster vaccine ranged from 50% to 75%, as shown in Table 2. In summary, individuals boosted with mRNA-1273 possessed the highest persistence of total RBD Ig and anti-RBD IgG among all booster groups, followed by half-dose mRNA-1273, half-dose BNT162b2, BNT162b2, AZD1222, and BBIBP, respectively.

**Table 2.** Measurement of GMTs and GMRs (with 95% confidence intervals) of total RBD Ig (U/mL) and anti-RBD IgG (BAU/mL) compared between day 28, and day 90–120 in each vaccine regimen.

|  | **BBIBP** | **AZD1222** | **BNT162b2 (II)** | **Half BNT162b2** | **mRNA-1273** | **Half-dose mRNA-1273** |
| --- | --- | --- | --- | --- | --- | --- |
|  | **(n=24)** | **(n=28)** | **(n=25)** | **(n=37)** | **(n=39)** | **(n=34)** |
| **SARS-CoV-2 total RBD Ig, (U/mL), GMT (95% CI)** |  |  |  |  |  |  |
| Day 28 | 1231 (891-1699) | 10766 (8235-14074) | 21240 (16478-27378) | 22345 (18596-26850) | 36845 (29827-45513) | 28087 (24041-32813) |
| Day 90 | 620 (425-904) | 3984 (3047-5209) | 5514 (4045-7517) | 9722 (7937-11909) | 10853 (8861-13292) | 10587 (8724-12848) |
| Day 90–120/28 ratio (GMR) | 0.50 (0.41-0.61) | 0.39 (0.33-0.45) | 0.26 (0.22-0.30) | 0.44 (0.39-0.49) | 0.30 (0.27-0.32) | 0.38 (0.34-0.42) |
| *P*-value | <0.001 | <0.001 | <0.001 | <0.001 | <0.001 | <0.001 |
| (Between D28 and D90–120) |  |  |  |  |  |  |
| %Reduction | 50.4% | 70.0% | 74.0% | 56.5% | 69.8% | 62.0% |
| **SARS-CoV-2 anti-RBD IgG, (BAU/mL), GMT (95% CI)** |  |  |  |  |  |  |
| Day 28 | 175 (127-241) | 1618 (1256-2082) | 3708 (2984-4607) | 4351 (3682-5141) | 7264 (6056-8714) | 5636 (4783-6641) |
| Day 90 | 84.9 (59.4-121) | 623 (481-808) | 917 (690-1219) | 1743 (1405-2162) | 1942 (1594-2365) | 1985 (1602-2459) |
| Day 90–120/28 ratio (GMR) | 0.49 (0.43-0.55) | 0.39 (0.32-0.45) | 0.25 (0.21-0.29) | 0.40 (0.36-0.45) | 0.27 (0.24-0.31) | 0.35 (0.31-0.40) |
| *P*-value | <0.001 | <0.001 | <0.001 | <0.001 | <0.001 | <0.001 |
| (Between D28 and D90–120) |  |  |  |  |  |  |
| %Reduction | 51.5% | 61.5% | 75.3% | 59.9% | 73.3% | 64.8% |

### 3.3 Surrogate virus neutralization-specific variants SARS-CoV-2 Wuhan, Omicron BA.2, and BA.4/5

A subset of 120 samples (20 samples/group) from day 90 to 120 was evaluated for surrogate neutralization activity against wild-type SARS-CoV-2, Omicron BA.2, and BA.5 subvariants. Most participants (85 to 100%) who had received the viral vector or the mRNA booster were seropositive for the wild type and (65% to 100%) for Omicron BA.2 and BA.4/5 subvariants. In contrast, only 61% and 20% to 40% of participants who received BBIBP booster presented seropositivity for wild type and Omicron BA.2 or BA.4/5 subvariants, respectively (Supplementary Table1). The median %inhibition of NAbs against the wild type remained higher than 95% for the viral vector and mRNA booster groups (Figure 2A). However, the median %inhibition of NAbs was substantially lower against the Omicron BA.2 subvariant at 26.8% in BBIBP, 60.3% in AZD1222, 60.0% in BNT162b2, and 77.7% in the half-dose BNT162b2, 75.4% in mRNA-1273, and 82.9% in the half-dose mRNA-1273 group (Figure 2B). Moreover, a reduction of %inhibition was also observed against the BA.4/5 subvariants with 15.6%, 50.2%, 45.6%, 80.5%, 67.5%, and 79.6% in the six groups, respectively (Figure 2C). In summary, the median %inhibition of neutralizing activity against the Omicron variants BA.2 and BA.5, was lower than that observed against the wild type variant. While the median %inhibition of neutralizing activity against the BA.5 was 5% to 15% lower compared to that against the BA.2 subvariant among all booster groups.

**Figure 2:**
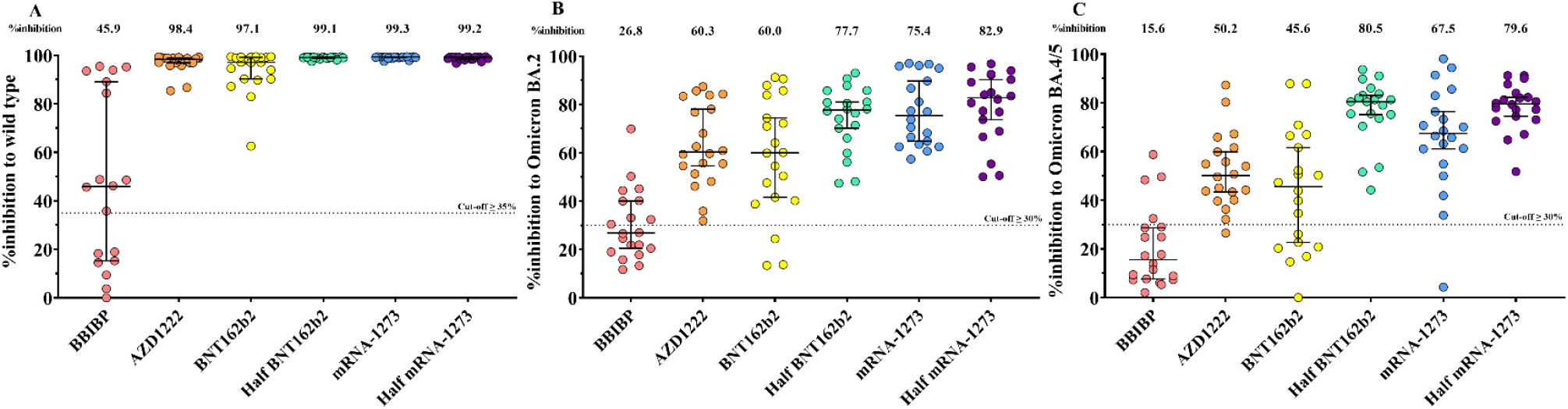
Neutralizing activities against wild type, Omicron BA.2, and BA.4/5 variants at day 90 to 120 post-boosted using a surrogate virus neutralization test. A subset of samples (n=20/group) from BBIBP, AZD1222, BNT162b, half-dose BNT162b2, mRNA-1273, and half-dose mRNA-1273 groups was randomly selected to test for sVNT. **(A)** Neutralizing activities against the SARS-CoV-2 wild type are shown as %inhibition and **(B)** Neutralizing activities against the SARS-CoV-2 Omicron variant (BA.2) are shown as %inhibition **(C)** Neutralizing activities against the SARS-CoV-2 Omicron variant (BA.4/5) are shown as %inhibition. Median values with interquartile ranges (IQRs) are denoted as horizontal bars. The dotted lines designate cutoff values at 35% for wild type and 30% for Omicron BA.2 and BA.5 subvariants.

### 3.4 Focus reduction neutralization test against the BA.1 and BA.2 variants

A subset of 20 samples from the AZD1222, BNT162b2, and mRNA-1273 groups on day 28 and day 90 to 120 was determined in the live virus focus reduction neutralization test (the BBIBP and the half-dose group were not included). Most of the participants (95% to 100%) on heterologous mRNA and viral vector booster schedules showed detectable NAbs on day 90 to 120. Previously, NAb GMTs against BA.1 were 243 for AZD1222, 313 for BNT162b2, and 645 for the mRNA-1273 groups at 28 days after the booster dose (Assawakosri et al., 2022b). Subsequently, the GMT of NAbs against BA.1 significantly decreased to 60.1, 82.6, and 133 in the three groups, respectively (Figure 3). While the NAbs GMTs against BA.2 were 340, 449, and 1035 in each group, which significantly dropped to 76.5, 70.5, and 229 in the AZD1222, BNT161b2 and mRNA-1273 groups, respectively. In comparison, the GMR D90–120/28 ratio of NAbs against BA.1 was approximately equal to the GMR D90–120/28 ratio of NAbs against BA.2. The percentage reduction among the three vaccines ranged between 70% and 85%, as shown in Supplementary Table 2. In accordance with those binding antibody levels, this finding demonstrated that the neutralizing antibody significantly decreased on day 90 to 120 compared to day 28 after the booster dose.

**Figure 3.**
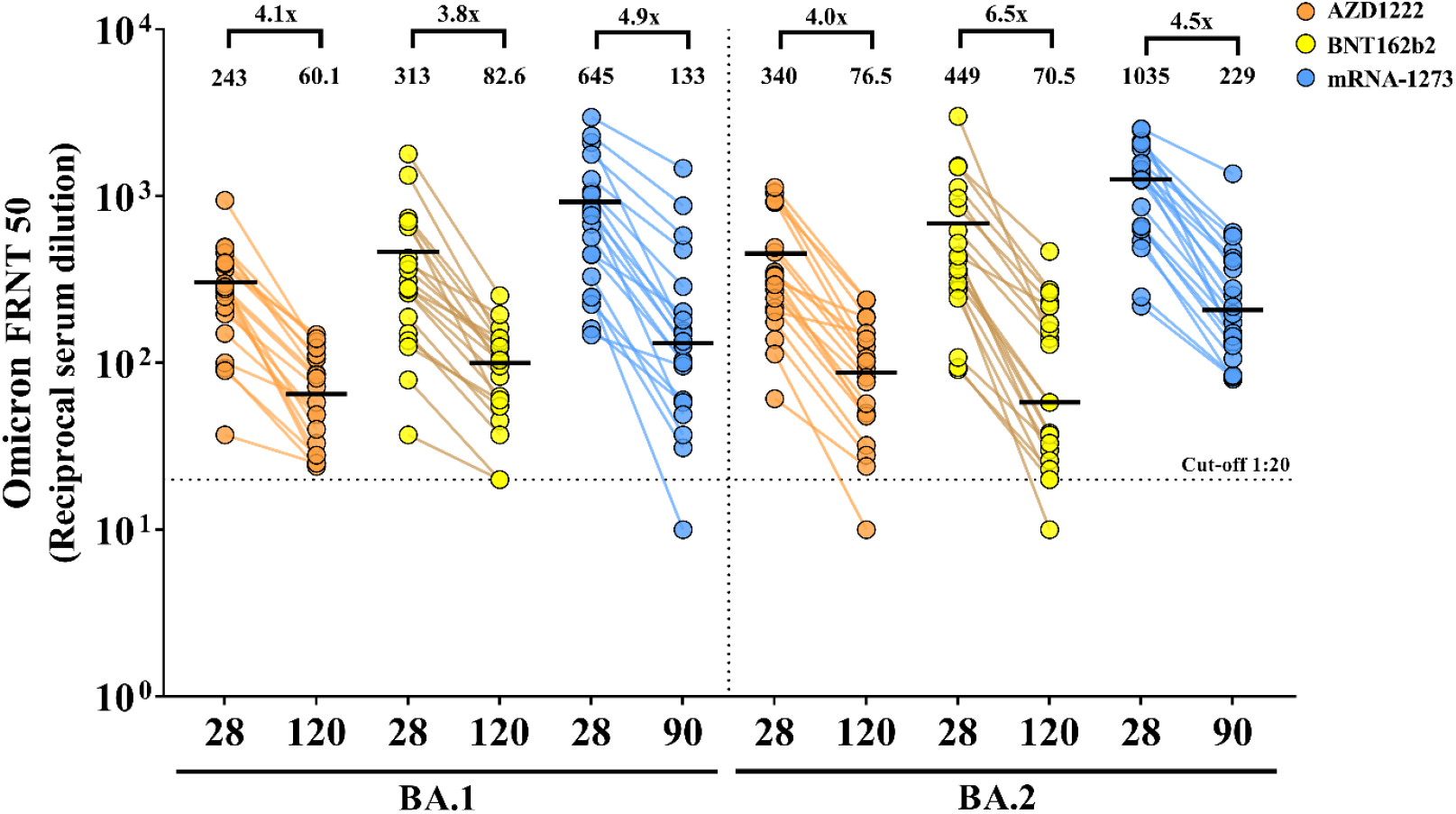
Comparison of neutralizing antibody titers against Omicron BA.1 and BA.2 between day 28 and day 90 to 120 after booster vaccination. A subset of samples (n=20/group) from AZD1222, BNT162b, and mRNA-1273 groups was randomly selected for testing of the focus reduction neutralization test (FRNT50) against SARS-CoV-2 Omicron BA.1 and BA.2 subvariant. Each data point represents an individual who received a booster vaccine, including the viral vector vaccine, AZD1222 (orange), the mRNA vaccine, BNT162b2 (yellow), or mRNA-1273 (blue). The error bars present GMT. Values below the limit were substituted with a titer of 10. The dotted lines designate cutoff values at 1:20.

## 4. Discussion

In this study, we determined the persistence levels of binding antibodies and NAbs against the RBD of SARS-CoV-2 on day 90 to 120 after implementing COVID-19 booster dose vaccines that included BBIBP, AZD1222, BNT161b2 (full- or half-dose), and mRNA-1273 (full- or half-dose) in individuals primed with two doses of CoronaVac. Our findings revealed that boosting with the heterologous vaccine elicited high levels of total RBD Ig, anti-RBD IgG, and NAbs at 28 days following booster vaccination. However, the antibody response substantially waned within three to four months after the booster dose, with a different pattern between each booster vaccine platforms. Additionally, consistent with the previous study in the United Kingdom, anti-spike IgG significantly decreased on day 84 after booster dose in participants who had previously been vaccinated with two doses of AZD1222 (Liu et al., 2022).

Total RBD Ig and anti-RBD IgG levels among all heterologous mRNA and viral vector booster vaccines highly remained on day 90 to 120 after vaccination. Conversely, binding antibody levels were lower in the BBIBP booster group. Among them, mRNA-1273 (full- and half-dose) demonstrated the highest level of total RBD Ig and anti-RBD IgG, while the decay rate of the mRNA-1273 booster was faster than that of the AZD1222 and BBIBP booster. Moreover, the decay rate of the binding antibodies was comparable between the full-dose and half-dose mRNA-1273 boosters. In agreement with the previous study of participants vaccinated with two doses of mRNA-1273 plus mRNA-1273 booster, the half-dose mRNA-1273 booster provided an antibody response similar to that of the full-dose booster (Choi et al., 2021). These results suggest that the half-dose mRNA-1273 is interchangeable with the full-dose mRNA-1273 with no difference in decay rate.

Several studies have demonstrated that neutralizing antibody level is correlated with vaccine-induced immune protection against SARS-CoV-2 symptomatic disease (Earle et al., 2021; Favresse et al., 2022; Khoury et al., 2021; Koch et al., 2021). Therefore, a higher level of NAb may represent a higher percentage of vaccine efficacy against severe infection. After 90 to 120 days post-vaccinated, we observed a high seropositive rate of NAb against the Omicron variant among participants who received heterologous mRNA and viral vector booster vaccines, especially mRNA-1273 vaccines. In line with a study in China that evaluated the use of the mRNA RQ3013 vaccine as the booster dose, mRNA booster provided the highest neutralizing antibodies against the omicron variant compared to other COVID-19 platforms (Zhang et al., 2022). In addition, we observed a comparable level of neutralizing antibodies against the BA.1 and the BA.2 subvariants. In accordance with those observed in the previous study, the neutralizing antibody titers against the BA.2 subvariant were approximately equal to the BA.1 subvariant and 1.7 times higher in participants with a homologous BNT162b2 booster (Chen et al., 2022; Kurhade et al., 2022).

The booster consisting of a heterologous mRNA and viral vector vaccine showed a higher seropositivity rate of neutralizing activity against BA.4/5, while the BBIBP booster achieved only 20% seropositivity. Similar to the study of three-dose CoronaVac and two-dose CoronaVac plus BNT162b2, the seropositive number of Plaque neutralizing antibody (PRNT50) against BA.4 and BA.5 was only 10% in homologous three-dose CoronaVac compared to 90–100% in a heterologous booster (Cheng et al., 2022). The results showed that the NAbs induced by heterologous mRNA and viral vector as a third dose vaccination had broad neutralizing activity against several subvariants of SARS-CoV-2. Consistent with a previous study, after the homologous mRNA booster, the percentage of somatic hypermutated memory B cells increased and indicated B cell affinity maturation (Paschold et al., 2022). In comparison, the median %inhibition of neutralizing activity against Omicron BA.2 and BA.4/5 showed a significantly lower susceptibility than against wild type. Whereas the median %inhibition of neutralizing activity against BA.4/5 also demonstrated a lower trend than that of BA.2. Consistent with the 50% neutralization titers against BA.4/5 data from homologous mRNA booster, BA.4/5 had lower neutralizing antibody titers than the BA.2.12.1 and D614G variants (Qu et al., 2022). These results indicated that the BA.4/5 subvariant exerts a higher vaccine-induced immune evasion and BA.4/5 certainly has overtaken other circulating subvariants.

Our findings showed that NAbs titers against the Omicron subvariant induced by the third dose booster waned over time, despite the fact that most participants had detectable NAbs on day 90 to 120 after the booster dose. According to real-world VE data from Brazil, the estimated VE against symptomatic infection by the Omicron variant in participants who received a primary series of CoronaVac plus the BNT162b2 booster significantly decreased from 63.6% at day 14– 30 to 1.7% at day 120 or more after the booster dose (Cerqueira-Silva et al., 2022a). Another study showed a decrease in VE from 57.3% at day 8–59 to 15.7% at day ≥120 after BNT162b2 booster, while the VE in the homologous CoronaVac booster was only 8.1% at day 8–59 and decreased to -24.8% at day ≥120 post-boosted (Ranzani et al., 2022). However, the estimated VE against hospitalization and death remained highly effective, with 84.1% at day 120 or more after the booster dose. These results suggested that the third dose mRNA vaccine still achieved protection against severe disease, although it showed less protection against infection. However, individuals with inactivated boosters may require additional booster doses with other vaccine platforms due to inadequate serological immune response and protection.

Our study has several limitations. First, due to the extension of the present study from the previous study cohort, almost 44% of all participants were lost to follow-up during the extended period at four-month follow-up timepoint. Second, the surrogate virus neutralization assay technique reached the upper detection limit for the wild type. Therefore, this method could not determine the exact NAb level. Lastly, data on the cellular immunity profile was not investigated. Data on the live virus neutralization test against other newly emerged BA.4/5 or BA.2.75 will be of great interest for further exploration.

## 5. Conclusion

At 90 to 120 days after third dose vaccination with the viral vector and mRNA in CoronaVac-primed individuals demonstrated high detectable rate of humoral immune responses both binding, and neutralizing antibodies against emerging Omicron BA.1, BA.2 and BA.4/5 SARS-CoV-2 subvariants. Nonetheless, real-world data has demonstrated a low percentage of VE against COVID-19 infection for individuals receiving any types of booster doses. To maintain the immune response against COVID-19 infection, a fourth booster dose is recommended for all groups, especially those who received the inactivated vaccine as the third dose booster.

## Data Availability

All data produced in the present work are contained in the manuscript

## Declaration

### Author contribution statement

Suvichada Assawakosri: Conceived and designed the experiments; Performed the experiments; Analyzed and interpreted the data; Wrote the paper.

Sitthichai Kanokudom, Nungruthai Suntronwong, Jira Chansaenroj: Performed the experiments; Analyzed and interpreted the data.

Chompoonut Auphimai, Pornjarim Nilyanimit, Preeyaporn Vichaiwattana, Thanunrat Thongmee, Thaneeya Duangchinda, Warangkana Chantima, Pattarakul Pakchotanon Donchida Srimuan, Thaksaporn Thatsanatorn, Sirapa Klinfueng: Performed the experiments.

Natthinee Sudhinaraset, Nasamon Wanlapakorn, Juthathip Mongkolsapaya, Sittisak Honsawek, Yong Poovorawan: Conceived and designed the experiments; Contributed reagents, materials, analysis tools or data.

## Acknowledgments

The authors are grateful to the Center of Excellence in Clinical Virology staffs, all participants, BJC Big C Foundation and MK restaurant group public Co., Ltd. for helping and supporting this project. We also thank the Ministry of Public Health, the Chulabhorn Royal Academy, and Zullig pharma for providing the vaccines for this study.

