## Supplementary material for "Immunogenicity and durability against Omicron BA.1, BA.2 and BA.4/5 variants at 3 to 4 months after a heterologous COVID-19 booster vaccine in healthy adults with a two-doses CoronaVac vaccination": Supplement.docx

**Supplementary Table 1.** The percentage inhibition against SARS-CoV-2 Wild-type, Omicron BA.2 and BA.4/5 subvariants on day 90 to 120 after booster vaccination.

| **Group**  **(n=20/group)** | **Wild type (EuroImmun®)** | | **Omicron BA.2 subvariant (GenScript®)** | | **Omicron BA.4/5 subvariant (GenScript®)** | |
| --- | --- | --- | --- | --- | --- | --- |
|  | **%Inhibition**  **median (IQR)** | **%Seropositive** | **%Inhibition**  **median (IQR)** | **%Seropositive** | **%Inhibition**  **median (IQR)** | **%Seropositive** |
| **BBIBP** | 45.9 (15.1-90.2) | 11/18 (61.1%) | 26.8 (19.4-40.2) | 8/20 (40.0%) | 15.6 (7.48-28.9) | 4/20 (20.0%) |
| **AZD1222** | 98.4 (96.8-98.9) | 20/20 (100%) | 60.3 (52.1-81.5) | 20/20 (100%) | 50.2 (40.9-61.2) | 19/20 (95.0%) |
| **BNT162b2** | 97.1 (90.2-99.2) | 20/20 (100%) | 60.0 (40.6-81.5) | 17/20 (85.0%) | 45.6 (21.3-65.3) | 13/20 (65.0%) |
| **Half-BNT162b2** | 99.1 (98.9-99.2) | 20/20 (100%) | 77.7 (67.1-84.5) | 20/20 (100%) | 80.5 (74.0-83.6) | 20/20 (100%) |
| **mRNA-1273** | 99.3 (99.0-99.3) | 20/20 (100%) | 75.4 (63.8-93.7) | 20/20 (100%) | 67.5 (56.5-81.4) | 19/20 (95.0%) |
| **Half-mRNA-1273** | 99.2 (98.4-99.3) | 20/20 (100%) | 82.9 (70.1-91.8) | 20/20 (100%) | 79.6 (73.5-83.6) | 20/20 (100%) |

**Supplementary Table 2** Value and statistical analysis of GMTs and GMRs (with 95% confidence intervals) of FRNT50 (BA.1 and BA.2) compared between day 28 and day 90 to 120 in AZD1222, BNT162b2, and mRNA-1273 vaccine regimen.

|  | AZD1222 (n=20) | BNT162b2 (n=20) | mRNA-1273 (n=20) |
| --- | --- | --- | --- |
| FRNT50 for BA.1 (Reciprocal serum dilution), GMT (95% CI) | | | |
| Day 28 | 243 (172-345) | 313 (202-486) | 645 (424-981) |
| Day 90 | 60.1 (44.4-81.3) | 82.6 (59.6-114) | 133 (76.0-232) |
| Day 90–120/28 ratio (GMR) | 0.25 (0.19-0.32) | 0.26 (0.21-0.34) | 0.21 (0.15-0.29) |
| *P*-value  (Between D28 and D90 to 120) | <0.001 | <0.001 | <0.001 |
| %Seropositive on day 90 to 120 | 20/20 (100%) | 20/20 (100%) | 19/20 (95.0%) |
| Fold-decreased | 4.1 | 3.8 | 4.9 |
| % Reduction | 75.3% | 73.6% | 79.4% |
| FRNT50 for BA.2 (Reciprocal serum dilution), GMT (95% CI) | | | |
| Day 28 | 340 (233-495) | 449 (289-696) | 1035 (740-1449) |
| Day 90 | 76.5 (51.6-114) | 70.5 (41.9-119) | 229 (160-329) |
| Day 90–120/28 ratio (GMR) | 0.23 (0.18-0.29) | 0.16 (0.11-0.22) | 0.22 (0.18-0.28) |
| *P*-value  (Between D28 and D90 to 120) | <0.001 | <0.001 | <0.001 |
| %Seropositive on days 90 to 120 | 19/20 (95.0%) | 19/20 (95.0%) | 20/20 (100%) |
| Fold-decreased | 4.4 | 6.4 | 4.5 |
| % Reduction | 77.5% | 84.3% | 77.9% |
